# Prospective validation of diffusion-weighted MRI as a biomarker of tumor response and oncologic outcomes in head and neck cancer: Results from an observational biomarker pre-qualification study

**DOI:** 10.1101/2022.04.18.22273782

**Authors:** Joint Head and Neck Radiotherapy-MRI Development Cooperative, Abdallah S. R. Mohamed, Abdelrahman Abusaif, Renjie He, Kareem Wahid, Vivian Salama, Sara Youssef, Brigid A. McDonald, Mohamed Naser, Yao Ding, Travis C. Salzillo, Moamen A. AboBakr, Jihong Wang, Stephen Y. Lai, Clifton D. Fuller

## Abstract

**Purpose/Objective(s):** To determine diffusion-weighted imaging (DWI) MRI parameters associated with tumor response and oncologic outcomes in head and neck (HNC) patients treated with definitive radiation therapy (RT).

**Materials/Methods:** Eighty-six HNC patients enrolled in an active prospective imaging study at The University of Texas MD Anderson Cancer Center were included in the analysis. Patients had MRIs pre-, mid-, and post-RT completion. Inclusion criteria included adults with histologic evidence of malignant head and neck neoplasm indicated for curative-intent treatment with RT with/without chemotherapy, good performance status (ECOG score 0-2), and with no contraindications to MRI. Patients were scanned using a MAGNETOM Aera 1.5T MR scanner (Siemens Healthcare, Germany) with two large four-channel flex phased-array coils. We used fat-suppressed T2-weighted turbo spin echo sequences for tumor segmentation which were co-registered to respective DWIs for extraction of apparent diffusion coefficient (ADC) measurements. Treatment response was assessed at mid-RT and at 8-12 weeks post-RT using RECIST 1.1 criteria and was defined as: complete response (CR) vs. non-complete response (non-CR). Pre-RT ADC was correlated with RT response (CR vs. non-CR) at mid- and post-RT. The Mann-Whitney U test was used to compare ADC values between the mid-treatment CR group and the non-CR group. Recursive partitioning analysis (RPA) was performed to identify ADC threshold associated with relapse. Cox proportional hazards models were done for clinical vs. clinical and imaging parameters and internal validation was done using bootstrapping technique.

**Results:** Eighty-one patients were included in this analysis. Median follow-up was 31 months. Pre-treatment ADC was not correlated with tumor response or oncologic outcomes (P>0.05). For patients with post-RT CR, there was a significant increase in mean ADC at mid-RT compared to baseline ((1.8 ± 0.29) × 10^−3^ mm^2^/s *versus* (1.37 ± 0.22) × 10^− 3^ mm^2^/s, *p* < 0.0001), while patients with non-CR had no statistically significant increase (*p* >0.05). RPA identified GTV-P delta (Δ) ADC_mean_ <7% at mid-RT as the most significant parameter associated with worse LC and RFS (*p*=0.01). Univariable and multivariable analysis of prognostic outcomes showed that GTV-P ΔADC_mean_ at mid-RT ≥7% was significantly associated with better LC and RFS. The addition of ΔADC_mean_ significantly improved the c-indices of LC and RFS models compared with standard clinical variables (0.85 vs. 0.77 and 0.74 vs. 0.68 for LC and RFS, respectively, *p*<0.0001 for both).

**Conclusion:** ADC change at mid-RT is a strong predictor of oncologic outcomes in HNC patients. Patients with no significant increase of primary tumor site ADC at mid-RT relative to baseline values are at high risk of disease relapse. Multi-institutional data are needed for validation of our results.

## Introduction

Radiation therapy (RT) is a cornerstone of head and neck cancer (HNC) treatment both in the definitive (i.e., organ preserving) and adjuvant post-operative setting. The goal of RT is to maximize the dose to cancer cells while minimizing the dose to adjacent normal tissues. However, tumors have variable sensitivity to RT leading to different disease response rates.(1) Current RT dose and fractionation are largely driven by empirical data rather than tumor-specific information regarding potential radiosensitivity or radioresistance.(2-5) The ability to predict tumor response before and/or during the RT course can allow for the adaptation of RT doses and potentially achieve better treatment outcomes for patients.

Non-invasive imaging such as magnetic resonance imaging (MRI) can provide important information related to tumor characteristics and response to RT. The development of MRI correlates of RT response would be critical for implementing adaptive RT strategies that maximize therapeutic ratios. Specifically, patients with aggressive non-responsive tumors may require RT dose escalation (3, 5), while patients with radiosensitive tumors may benefit from dose de-escalation to spare normal tissues with equivalent tumor control.(4) This represents a significant unmet clinical need since patients with radiosensitive tumors are over-treated and patients with radio-resistant tumors are under-treated. A leading-edge solution to the anatomic adaptive therapy problem has been to integrate MRI into radiation delivery devices (e.g., MR-Linear accelerators).(6) The richer data of MRI compared with standard-of-care CT images enables computer-driven identification of tumors and normal tissues and allows radiation plans to be adapted on a daily basis with limited human intervention. (7, 8) Yet, gross anatomic changes represent only one dimension of patient response to RT. Having incorporated high-field MRI into delivery devices, there is now the potential to monitor the biologic changes within the patient using functional MRI sequences without excess radiation, contrast exposure, or excess burden on patients’ time.

The central hypothesis of this study is that quantitative MR diffusion-weighted imaging (DWI) can be used as a predictive biomarker of treatment response and oncologic outcomes in HNC. Functional changes in a tissue (e.g., a reduction in cellular density through RT-induced apoptosis) is reflected in an alteration in the detected diffusion measures, using a metric known as the apparent diffusion coefficient (ADC). The ADC component of DWI has been previously used to detect treatment response in HNC.(9-11) Specifically, DWI has been shown to predict response to induction chemotherapy(12, 13), RT(11, 13-20), and tumor recurrence(21). Preliminary data from a prospective trial at our institution(22), supported by other group’s data(13, 15-17, 20, 23, 24), has demonstrated that DWI was able to discriminate patients who will have a complete response at mid-RT. Additionally, recent data from our group demonstrated that early tumor regression rate ≥25% at fraction 15 (i.e., mid-RT) in HNC patients is associated with better local control and overall survival.(25) These low-risk patients represent suitable candidates for RT dose de-escalation if dose could be coupled to a quantitative marker of tumor response probability (i.e., ADC). However, these findings remain to be validated in larger prospective studies with more mature follow-up data to correlate with oncologic outcomes and overall survival. To this end, we aim to determine DWI parameters associated with tumor response and oncologic outcomes in a prospective cohort of HNC patients treated with definitive RT.

## Methods

### Patient selection

HNC patients enrolled in an active prospective imaging study (NCT03145077) from January 2017 to March 2021 were included after institutional-review board approval and study-specific informed consent. Patients in this cohort had MRIs at pre-RT, mid-RT, and post-RT. Inclusion criteria were adult patients with histologic evidence of malignant head and neck neoplasm obtained from the primary tumor or metastatic lymph node; indicated for curative-intent treatment with radiotherapy with or without chemotherapy (induction or concurrent); good performance status (ECOG score 0-2); and with no contraindications to MRI. Patients evaluated in this study received RT using standard daily fractionation for a period of 6-7 weeks. Tumor staging was based on clinical imaging consisting of contrast (CE) CT prior to treatment initiation using current AJCC 8th edition staging criteria.

### MR Imaging

All patients enrolled in the study had imaging acquired using individualized immobilization devices. Head immobilization was performed to decrease motion artifacts during the imaging study, according to the methodology presented previously by our group.(26) Patients were scanned using a MAGNETOM Aera 1.5T MR scanner (Siemens Healthcare, Erlangen, Germany) with two large four-channel flex phased-array coils. After the scout scan, an anatomic T2-weighted (T2w) fast spin-echo sequence (TR/TE = 4.8 s/80 ms; echo train length = 15, pixel bandwidth = 300 Hz, slice thickness= 2 mm, matrix= 512 × 512) was performed. 120 axial slices with a field of view (FOV) of 25.6 cm were selected to cover the primary tumor and neck nodes. Acquisition parameters for DWI MRI were multi shot radial turbo spin-echo (i.e., BLADE), axial acquisition; TR = 6.5 s; TE = 50 ms; pixel bandwidth = 1220 Hz; FOV = 25.6 × 25.6 cm^2^; echo train length = 15; EPI factor = 7, acquisition matrix = 128 × 128; voxel size = 1 × 1 × 2 mm^3^; 24 contiguous slices; two b-values = 0 and 800 (sec/mm^2^) for each orthogonal diffusion direction; number of averages = 2 for b=0 and 8 for b=800. DWI acquisition of patients scanned after 2018 was performed with multi-shot spin-echo echo-planar imaging (i.e. readout segmentation of long variable echo-trains, RESOLVE), axial acquisition; TR = 3.5 s; TE = 65 ms; pixel bandwidth = 780 Hz; FOV = 25.6 × 25.6 cm^2^; acquisition matrix = 128 × 128; slice thickness = 4 mm; reconstruction voxel size = 1 × 1 × 2 mm^3^, 48 contiguous slices; two b-values = 0 and 800 (sec/mm^2^) for each orthogonal diffusion direction; number of averages = 2 for b=0 and 8 for b=800. ADC maps were subsequently autogenerated using a scanner-specific on-line software during image generation. RESOLVE was selected because of shorter scan time (3:30 vs. 7:03 minutes for BLADE) and relatively higher signal-to-noise ratio. Our quality assurance study using phantom, volunteer, and patient images showed that both methods display similar ADC values with no differences in repeatability studies.(27)

### Image Segmentation/Registration

The regions of interest (ROIs) for the primary gross tumor volume (GTV-P) and the nodal gross tumor volume (GTV-N) were manually segmented by an expert radiation oncologist (ASRM) using the pre-RT T2w images. Deformable image registration (DIR) was used to register MR sequences at different time points (i.e., baseline and mid-RT) using the benchmarked commercially available image registration software (Velocity AI, version 3.0.1, Atlanta, GA). All baseline GTV-P ROIs were then propagated to the mid-RT T2w images (i.e., mid-RT GTV-P) which represent the same three-dimensional (3D) volume of the original GTV-P on mid-RT images and include both responding and non-responding voxels. This was followed by quality assurance (QA) review and manual editing whenever needed to exclude air gaps or non-anatomically relevant parts in case of massive tumor shrinkage. Residual GTV-N ROIs, on the other hand, were all manually segmented on mid-RT images. Subsequently, DWI images were co-registered with the corresponding T2w of each time point and finally all ROIs were propagated to extract corresponding ADC values. Additional ROIs were created on mid-RT images for patients with non-complete GTV-P response at mid-RT to assess DWI differences between responding and non-responding sub-volumes within the mid-RT GTV-P. The first sub-volume was labeled mid-RT GTV-P-RD which represents the residual disease and the second sub-volume was labeled mid-RT GTV-P-RS which represents the area of response. Figure 1 illustrates the workflow process for image registration and segmentation.

**Figure 1.**
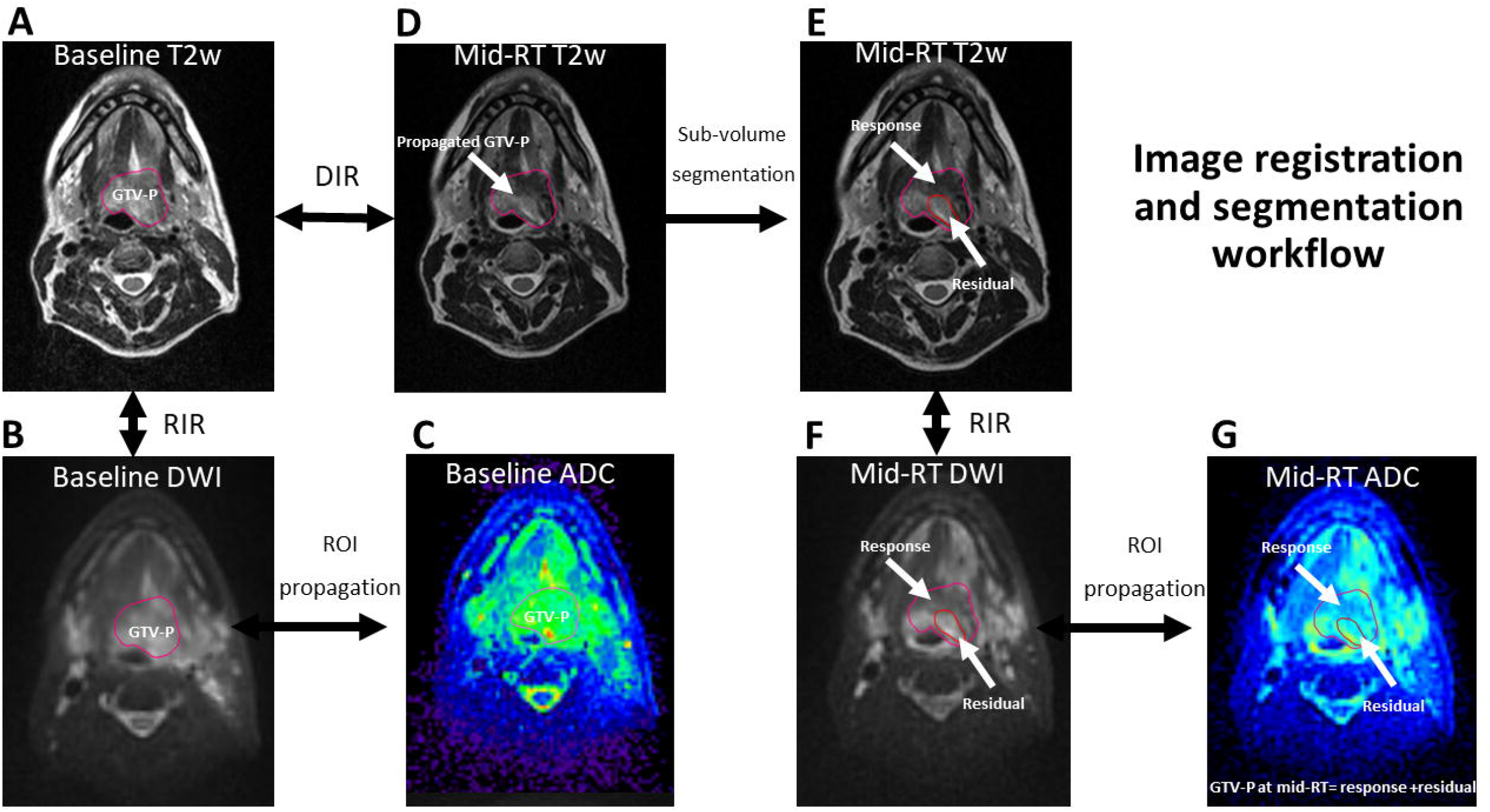
Illustration of the workflow process for image registration and segmentation in the study using an example of a patient with T4N1 tumor of the base of tongue. Panel (A) shows the GTV-P segmentation on baseline T2w MRI followed by rigid co-registration (RIR) and contour propagation to baseline DWI (B) and then ROI propagation to corresponding ADC map (C). Panel (D) shows mid-RT T2w image with partial response. The image was co-registered to baseline T2w using deformable image registration (DIR) and baseline GTV-P was propagated. Subsequently, the residual and response sub-volumes were segmented (E), then contours were propagated to mid-RT DWI after RIR (F), and finally to the corresponding mid-RT ADC map (G).

### Outcome definition

Treatment response was assessed at mid-RT and at 8-12 weeks post-RT using RECIST 1.1 criteria and was defined as: complete response (CR) vs. non-complete response (non-CR). All patients had complete physical examination, fiberoptic endoscopy, MRI, and CECT or FDG PET-CT performed 8-12 weeks after RT completion to assess the final treatment response. Oncologic outcomes included two-year local control (LC), regional control (RC), freedom from distant metastasis (FDM), recurrence-free survival, and overall survival.

### Statistical analysis

Continuous data were presented as mean ± standard deviation (SD), and categorical data were presented as proportions. The Kolmogorov–Smirnov test was used to assess the difference in baseline ADC in BLADE vs. RESOLVE DWI sequences. The ADC values for all voxels included in GTV-P and GTV-N ROIs were assessed by histogram analysis and the following parameters were extracted using in-house MATLAB script (MATLAB, MathWorks, MA, USA): ADC mean, 5th, 10th, 20th, 30th, 40^th^, 50th (i.e. median), 60th, 70th, 80th, 90th, 95^th^ percentile. Pre-RT ADC parameters were correlated with RT response (CR vs. non-CR) at mid- and post-RT time points using the non-parametric Mann-Whitney U test to compare ADC values between the mid-RT CR and non-CR groups. The non-parametric Wilcoxon signed-rank test was used to compare the mid-RT versus baseline ADC. Delta ADC (ΔADC) were calculated as the percent change of ADC relative to baseline value for each parameter 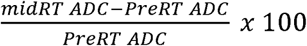. Delta volumetric changes for both GTV-P and GTV-N at mid-RT were also calculated and the non-parametric Spearman’s Rho test was used to determine the relationship between ΔADC and Δ volume changes. Recursive partitioning analysis (RPA) was performed to identify ΔADC threshold associated with relapse. Oncologic and survival endpoints were calculated using the Kaplan-Meier method and the statistical significance was determined using a p-value <0.05. Uni- and multi-variable analyses for oncologic and survival endpoints were performed using Cox regression. For multivariable analysis, we tested the impact of including the ADC parameter of choice compared with baseline models of standard clinical variables. We subsequently compared the new model using Bayesian information criteria (BIC).(28) A lower BIC indicates improved model performance and parsimony, using the BIC evidence grades presented by Raftery (29) with the posterior probability of superiority of a lower BIC model, where a BIC decrease of < 2 is considered “Weak” (representing a 50–75% posterior probability of being superior model), 2–6 denoted “Positive” (posterior probability of 75–95%), 6–10 as “Strong” (posterior probability of > 95%), and > 10, “Very strong” (posterior probability > 99%). In addition, Cox proportional hazards models were constructed using the scikit-survival package in Python version 3.9.7.(30) We initially constructed standard clinical models that include T stage, HPV status, and smoking status for LC prediction and AJCC stage 8th edition, age at diagnosis, and smoking status for RFS prediction. The selection of these clinical variables was based on the findings of our group’s large-scale HNC clinical models’ performance for survival endpoints prediction.(31) Subsequently, additive models that include the clinical parameters plus ΔADC were constructed to assess the potential additive value of the imaging parameter. Models were only constructed for patients with a GTV-P. Patients were split into training (85%) and testing (15%) sets through a bootstrap procedure (1000 iterations) for the internal validation and evaluation of constructed models. Mean and standard deviation of c-index values across all bootstrap iterations were reported for each model. Wilcoxon signed-rank tests were applied to compare clinical and additive models for each outcome. All other analyses were executed with JMP Pro version 15 software (SAS Institute, Cary, NC). The analysis and reporting of the results of this study adopted the reporting recommendations for tumor marker prognostic studies (REMARK) checklist.(32)

## Results

### Patients

Eighty-six patients were enrolled. Five patients were excluded from this analysis because of lack of visible GTVs after induction chemotherapy (n = 3) and loss to follow-up (n = 2) leaving a total of 81 patients in the final analysis. At pre-RT, 53 patients had both baseline GTV-P and GTV-N, 6 patients had baseline GTV-P without GTV-N, and 22 patients had GTV-N with no GTV-P (i.e., total GTV-P=59 and total GTV-N=74). Patients with no visible GTV-P at baseline had either carcinoma of neck nodes of unknown primary (CUP; n=12), tonsillectomy prior to RT (n=6), or CR to induction chemotherapy (n=4). The majority of patients were men (n=74, 93%) and the median age was 61 years (range 33-78). Most patients had human papillomavirus (HPV) positive disease (n= 73, 90%). A summary of patient demographic, disease, and treatment criteria is presented in Table 1.

**Table 1.**
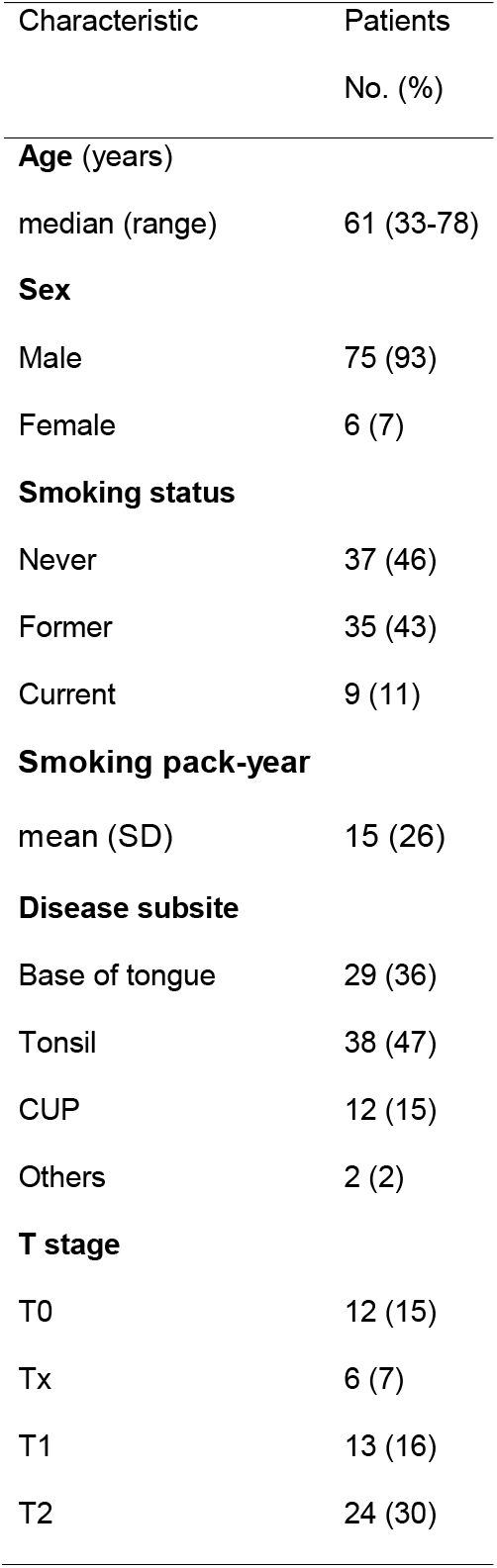
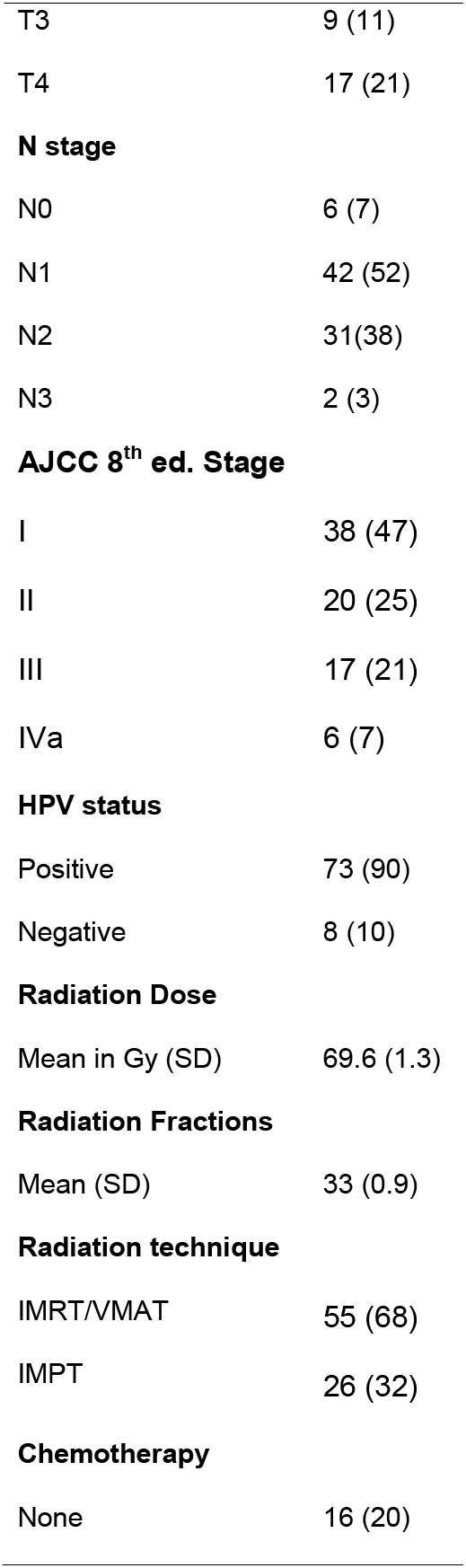
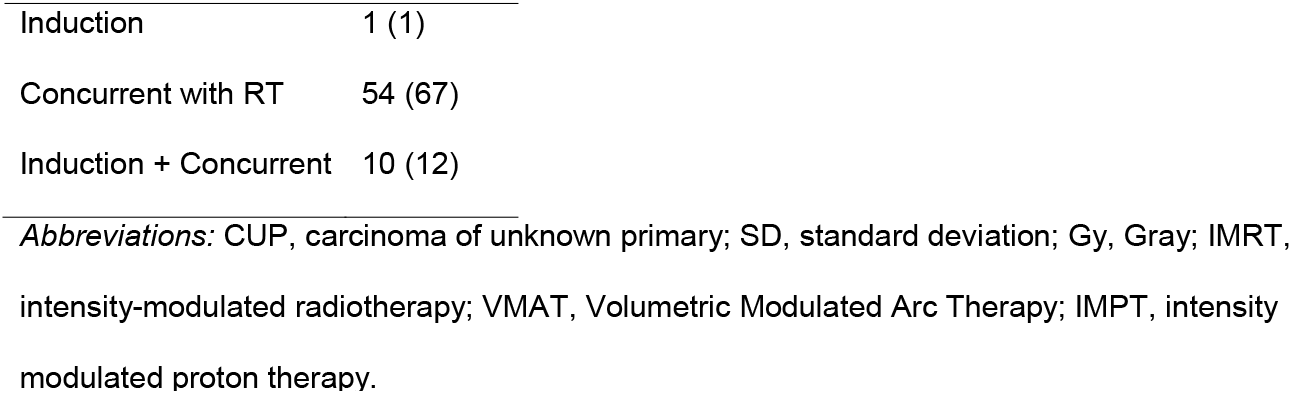
Patient demographic, disease, and treatment characteristics.

| Characteristic | Patients |
| --- | --- |
|  | No. (%) |
| **Age (years)** |  |
| median (range) | 61 (33-78) |
| **Sex** |  |
| Male | 75 (93) |
| Female | 6 (7) |
| **Smoking status** |  |
| Never | 37 (46) |
| Former | 35 (43) |
| Current | 9 (11) |
| **Smoking pack-year** |  |
| mean (SD) | 15 (26) |
| **Disease subsite** |  |
| Base of tongue | 29 (36) |
| Tonsil | 38 (47) |
| CUP | 12 (15) |
| Others | 2 (2) |
| **T stage** |  |
| T0 | 12 (15) |
| Tx | 6 (7) |
| T1 | 13 (16) |
| T2 | 24 (30) |
| T3 | 9 (11) |
| T4 | 17 (21) |
| **N stage** |  |
| N0 | 6 (7) |
| N1 | 42 (52) |
| N2 | 31(38) |
| N3 | 2 (3) |
| **AJCC 8<sup>th</sup> ed. Stage** |  |
| I | 38 (47) |
| II | 20 (25) |
| III | 17 (21) |
| IVa | 6 (7) |
| **HPV status** |  |
| Positive | 73 (90) |
| Negative | 8 (10) |
| **Radiation Dose** |  |
| Mean in Gy (SD) | 69.6 (1.3) |
| **Radiation Fractions** |  |
| Mean (SD) | 33 (0.9) |
| **Radiation technique** |  |
| IMRT/VMAT | 55 (68) |
| IMPT | 26 (32) |
| **Chemotherapy** |  |
| None | 16 (20) |
| Induction | 1 (1) |
| Concurrent with RT | 54 (67) |
| Induction + Concurrent | 10 (12) |

*Abbreviations:* CUP, carcinoma of unknown primary; SD, standard deviation; Gy, Gray; IMRT, intensity-modulated radiotherapy; VMAT, Volumetric Modulated Arc Therapy; IMPT, intensity modulated proton therapy.

### Treatment outcomes

For patients with GTV-P at baseline (n=59), 18 (31%) had mid-RT CR at the primary site which increased to 53 (90%) post-RT. Only 6 patients (10%) had persistent local disease as assessed by imaging at post-RT. Among the 6 patients, all had subsequent pathological confirmation of residual/recurrent disease. For patients with GTV-N at baseline (n=75), no patient had CR at the neck at mid-RT while 65 patients (87%) had CR as assessed by imaging at post-RT. Upon further pathological assessment, 6 out of 10 patients with non-CR at the neck had residual/recurrent disease while the reminder had necrotic non-active tissue.

The median follow-up time was 31 months (IQR, 18-38). The 2-year LC, RC, and FDM for the entire cohort were 91%, 92%, and 91%, respectively. The 2-year RFS and OS were 83% and 94%, respectively. The total number of recurrence events was 15 (18%). Two, three, and five patients had an isolated local, regional, and distant recurrence events, respectively. One, two, and two patients had combined local & distant, locoregional, and locoregional & distant recurrences, respectively.

### DWI correlates of outcomes

#### Baseline ADC parameters

Baseline mean, median, and different histogram percentile ADC values for BLADE vs. RESOLVE were not significantly different for both GTV-P and GTV-N ROIs (*p* >0.05 for both, Figure 2). There was no statistically significant correlation between pre-RT ADC parameters and CR at mid-RT and post-RT time points for GTV-P. Similarly, there was no significant correlation between pre-RT parameters and CR at post-RT for GTV-N (*p* >0.05 for all). Univariable analysis also did not show a significant correlation between pre-RT ADC parameters and all oncologic and survival endpoints.

**Figure 2.**
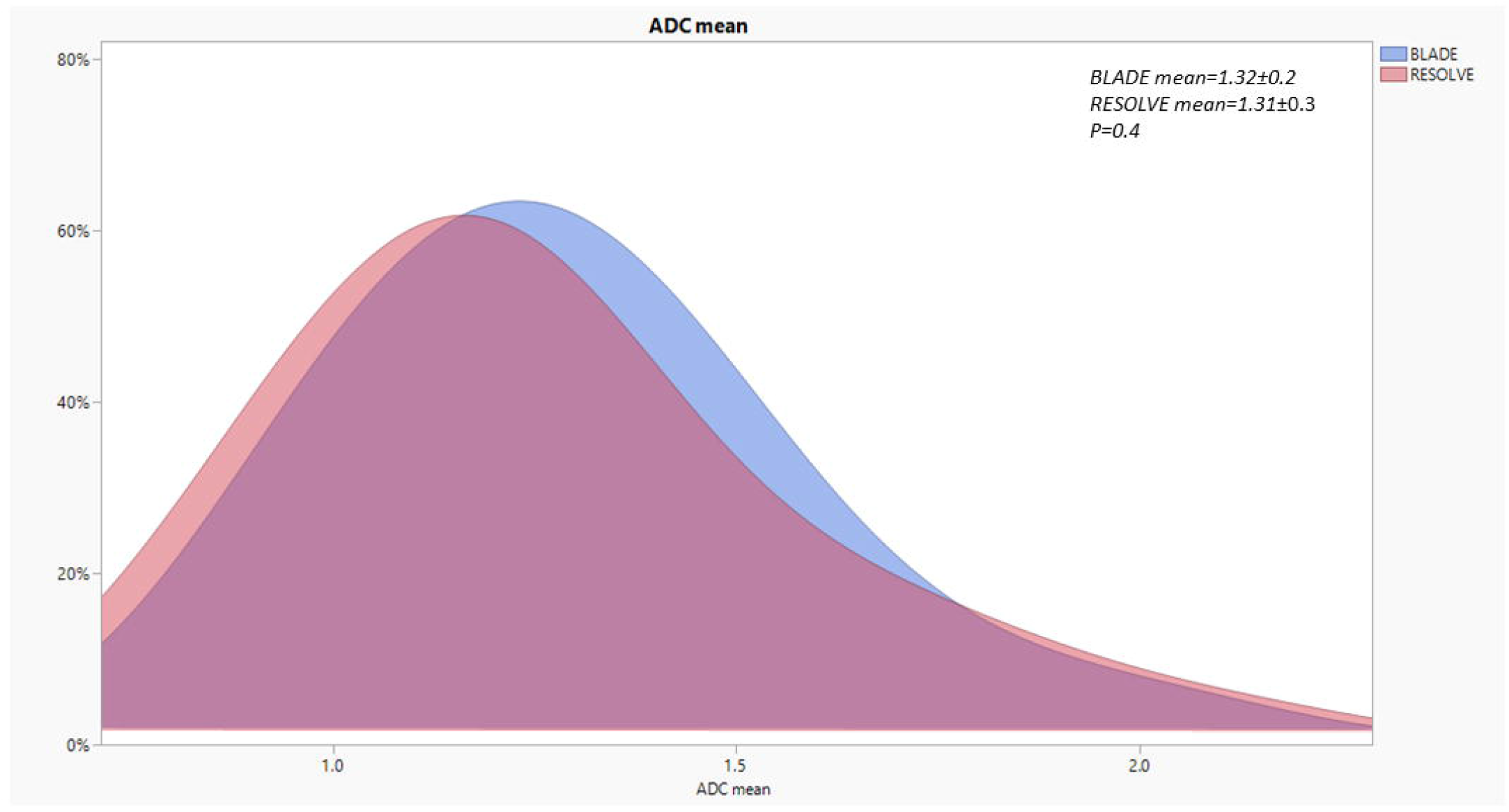
BLADE vs. RESOLVE histograms. Histogram illustration of the distribution of tumor and nodal volumes’ ADC mean at baseline using the BLADE vs. RESOLVE DWI acquisition methods in the study. The RESOLVE in pink is overlaid on BLADE in light blue. There were no statistically significant differences using the Kolmogorov–Smirnov test (*p*=0.4).

#### Mid-RT and delta ADC parameters

There was a statistically significant increase in all mid-RT GTV-P ADC parameters compared to baseline values (*p* <0.0001 for all, Table 2). Additionally, there was a statistically significant increase in all mid-RT GTV-N ADC parameters compared to baseline values (*p* <0.0001 for all, Table 2). For patients with CR of the primary tumor at the end of RT, there was a significant increase in GTV-P ADC_mean_ at mid-RT compared to baseline ((1.8 ± 0.29) × 10^− 3^ mm^2^/s *versus* (1.37 ± 0.22) × 10^−3^ mm^2^/s, *p* < 0.0001). On the other hand, patients with non-CR had no statistically significant increase in GTV-P ADC_mean_ (*p*>0.05). All other studied ADC parameters also had a significant increase at mid-RT for patients with CR of the primary tumor at the end of RT compared to non-CR. However, there was a significant increase in GTV-N ADC parameters at mid-RT for both patients with CR and non-CR at the end of RT.

**Table 2.** ADC parameter changes at mid-RT versus baseline values.

| ADC |  |  |  |  |  |  |  |
| --- | --- | --- | --- | --- | --- | --- | --- |
| parameter<br>($\times 10^{-3}$<br>$\text{mm}^2/\text{s}$) | End-RT<br>response | Baseline<br>GTV-P | Mid-RT<br>GTV-P | P value | Baseline<br>GTV-N | Mid-RT<br>GTV-N | P value |
| ADC Mean | CR | 1.37±0.2 | 1.8±0.3 | <0.0001 | 1.27±0.3 | 1.6±0.4 | <0.0001 |
| | Non-CR | 1.3±0.2 | 1.6±0.4 | 0.07 | 1.28±0.3 | 1.54±0.3 | 0.01 |
| ADC 5th<br>percentile | CR | 0.87±0.3 | 1.2±0.3 | <0.0001 | 0.76±0.2 | 1.04±0.3 | <0.0001 |
| | Non-CR | 1.1±0.6 | 1.3±0.5 | 0.4 | 1.02±0.4 | 1.19±0.3 | 0.2 |
| ADC 10th<br>percentile | CR | 0.97±0.3 | 1.32±0.3 | <0.0001 | 0.86±0.2 | 1.16±0.3 | <0.0001 |
| | Non-CR | 1.2±0.5 | 1.42±0.4 | 0.3 | 1.06±0.3 | 1.26±0.2 | 0.02 |
| ADC 20th<br>percentile | CR | 1.1±0.3 | 1.47±0.3 | <0.0001 | 0.97±0.3 | 1.3±0.4 | <0.0001 |
| | Non-CR | 1.2±0.4 | 1.47±0.3 | 0.4 | 1.11±0.3 | 1.34±0.2 | 0.01 |
| ADC 30th<br>percentile | CR | 1.17±0.3 | 1.58±0.3 | <0.0001 | 1.06±0.3 | 1.4±0.4 | <0.0001 |
| | Non-CR | 1.2±0.3 | 1.5±0.3 | 0.3 | 1.16±0.2 | 1.4±0.1 | 0.009 |
| ADC 40th | CR | 1.25±0.3 | 1.68±0.3 | <0.0001 | 1.14±0.3 | 1.5±0.4 | <0.0001 |
| percentile | Non-CR | 1.22±0.3 | 1.56±0.3 | 0. | 1.21±0.2 | 1.45±0.2 | 0.01 |
| ADC Median | CR | 1.35±0.2 | 1.8±0.3 | <0.0001 | 1.22±0.3 | 1.58±0.4 | <0.0001 |
|  | Non-CR | 1.25±0.2 | 1.6±0.2 | 0.07 | 1.25±0.3 | 1.51±0.2 | 0.009 |
| ADC 60th | CR | 1.42±0.3 | 1.9±0.4 | <0.0001 | 1.32±0.4 | 1.67±0.4 | <0.0001 |
| percentile | Non-CR | 1.3±0.2 | 1.68±0.5 | 0.1 | 1.3±0.3 | 1.57±0.3 | 0.009 |
| ADC 70th | CR | 1.52±0.3 | 1.95±0.4 | <0.0001 | 1.43±0.4 | 1.78±0.5 | <0.0001 |
| percentile | Non-CR | 1.36±0.3 | 1.7±0.6 | 0.1 | 1.36±0.4 | 1.64±0.4 | 0.009 |
| ADC 80th | CR | 1.6±0.3 | 2.1±0.4 | <0.0001 | 1.57±0.4 | 1.91±0.5 | <0.0001 |
| percentile | Non-CR | 1.4±0.3 | 1.8±0.7 | 0.07 | 1.45±0.4 | 1.72±0.5 | 0.01 |
| ADC 90th | CR | 1.79±0.4 | 2.22±0.4 | <0.0001 | 1.76±0.4 | 2.07±0.5 | <0.0001 |
| percentile | Non-CR | 1.52±0.4 | 1.99±0.8 | 0.03 | 1.57±0.5 | 1.84±0.6 | 0.1 |
| ADC 95th | CR | 1.9±0.4 | 2.36±0.5 | <0.0001 | 1.93±0.4 | 2.2±0.5 | <0.0001 |
| percentile | Non-CR | 1.61±0.6 | 2.09±0.9 | 0.03 | 1.65±0.7 | 1.97±0.8 | 0.1 |

RPA analysis identified GTV-P ΔADC_mean_ <7% at mid-RT as the most significant parameter associated with worse LC and RFS (*p* =0.01). The 2-Year LC and RFS for patients with ΔADC_mean_ <7% compared to patients with ≥7% at mid-RT were 48% and 42% versus 96% and 87%, respectively (*p* <0.0001 and 0.001, Figure 3). Δ GTV-N ADC parameters at mid-RT, however, were not significantly associated with any of the studied endpoints (P>0.05).

**Figure 3.**
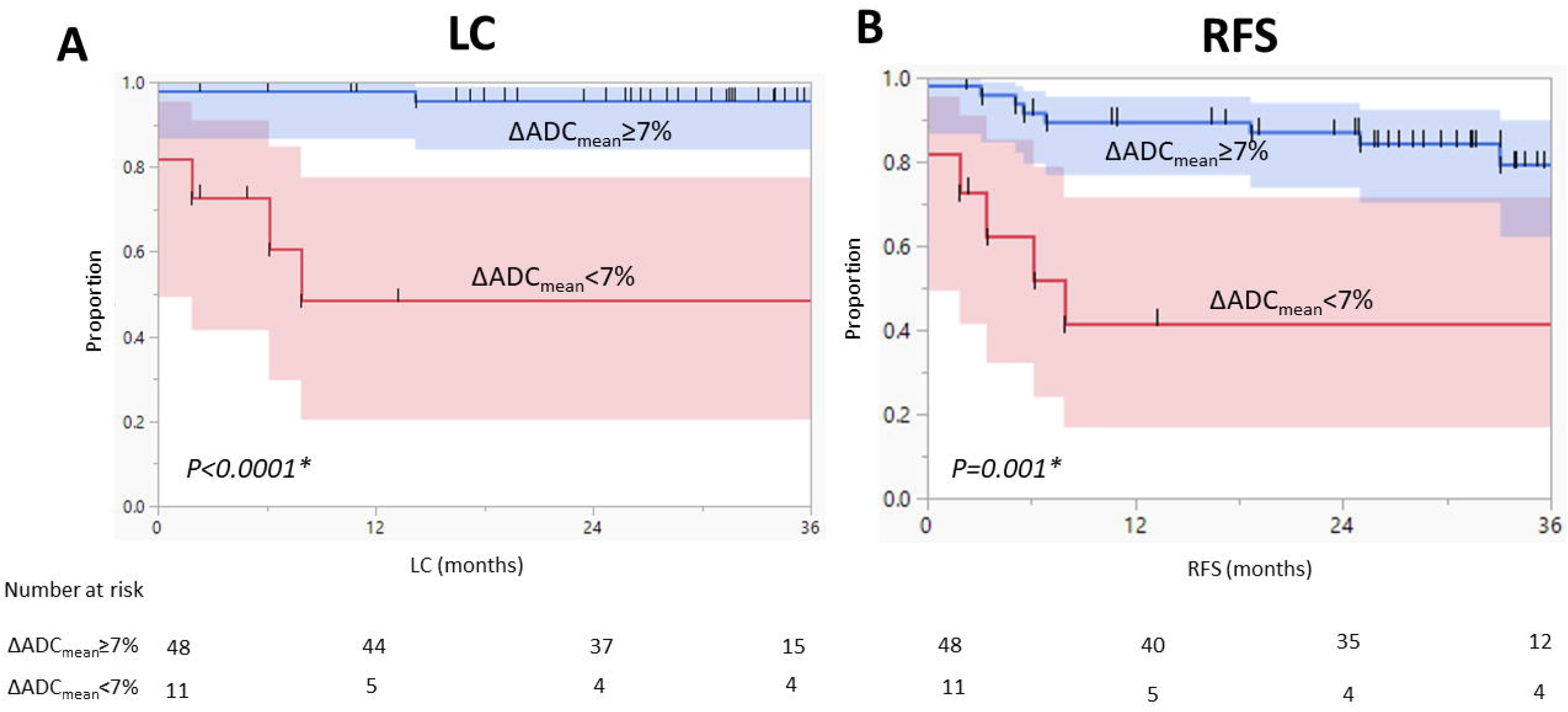
Kaplan–Meier curves calculated for patients with baseline GTV-P (n = 59) show better (A) local control (LC) and (B) recurrence-free survival (RFS) for patients with ≥7% ΔADC_mean_ at mid-RT. Shaded colors represent 95% confidence intervals, short vertical lines represent censored data, and asterisks indicate significant log-rank p values.

Univariable analysis of local control showed that GTV-P ΔADC_mean_ at mid-RT ≥7% was associated with improved LC (hazard ratio (HR), 0.06, 95% CI, 0.01-0.3, *p* =0.001). In a multivariable model that also included T-stage, smoking, and HPV status, GTV-P ΔADC_mean_ at mid-RT remained statistically significant (HR, 0.03, 95% CI, 0.01-0.6, *p* =0.02) and achieved a better model performance as assessed using BIC criteria (BIC decrease =19.8). After bootstrapping, the clinical LC model yielded a c-index of 0.77 ± 0.17 while the additive LC model (i.e., clinical + ΔADC_mean_) yielded a c-index of 0.85 ± 0.16 which was significantly better than the clinical model (*p* < 0.0001).

Moreover, univariable analysis of recurrence-free survival showed that GTV-P ΔADC_mean_ at mid-RT ≥7% was associated with improved RFS (HR, 0.2, 95% CI, 0.06-0.6, *p* =0.003). In a multivariable model that also included age, AJCC 8th edition stage (i.e., which is based on T-stage, N-stage, tumor site and HPV-status data), and smoking status, GTV-P ΔADC_mean_ at mid-RT remained statistically significant (HR, 0.3, 95% CI, 0.1-0.9, *p* =0.04) and also improved the model performance using BIC criteria (BIC decrease =8). After bootstrapping, the clinical RFS model yielded a c-index of 0.68 ± 0.23 while the additive RFS model yielded a c-index of 0.74 ± 0.22 which also was significantly better than the clinical models (*p* < 0.0001).

Similarly, a univariable analysis of overall survival showed that GTV-P ΔADC_mean_ at mid-RT ≥7% was associated with improved OS (HR, 0.2, 95% CI, 0.04-0.9, *p* =0.037). However, it was not statistically significant when added to a multivariable clinical model.

#### Volumetric analysis and ADC

There was a significant decrease in mid-RT residual tumor volumes for both GTV-P and GTV-N compared to baseline pre-RT volumes (3.5 vs. 11.1 mm^3^ for GTV-P and 7.4 vs. 11.8 mm^3^ for GTV-N, *p* <0.0001 for both). However, the mean Δ volume decrease at mid-RT was significantly higher in GTV-P compared with GTV-N (69% vs. 30%, *p* <0.0001). As shown in Figure 4, there was no statistically significant correlation of the Δ volume and ΔADC_mean_ for both GTV-P (Spearman’s Rho=-0.06, *p* =0.6) and GTV-N (Spearman’s Rho=-0.2, *p* =0.1). Δ volume changes were not significantly correlated with any endpoints (P>0.05). Only baseline GTV-P volume (i.e., a surrogate of T-stage) was significantly correlated with LC on univariable analysis (*p* =0.03).

**Figure 4.**
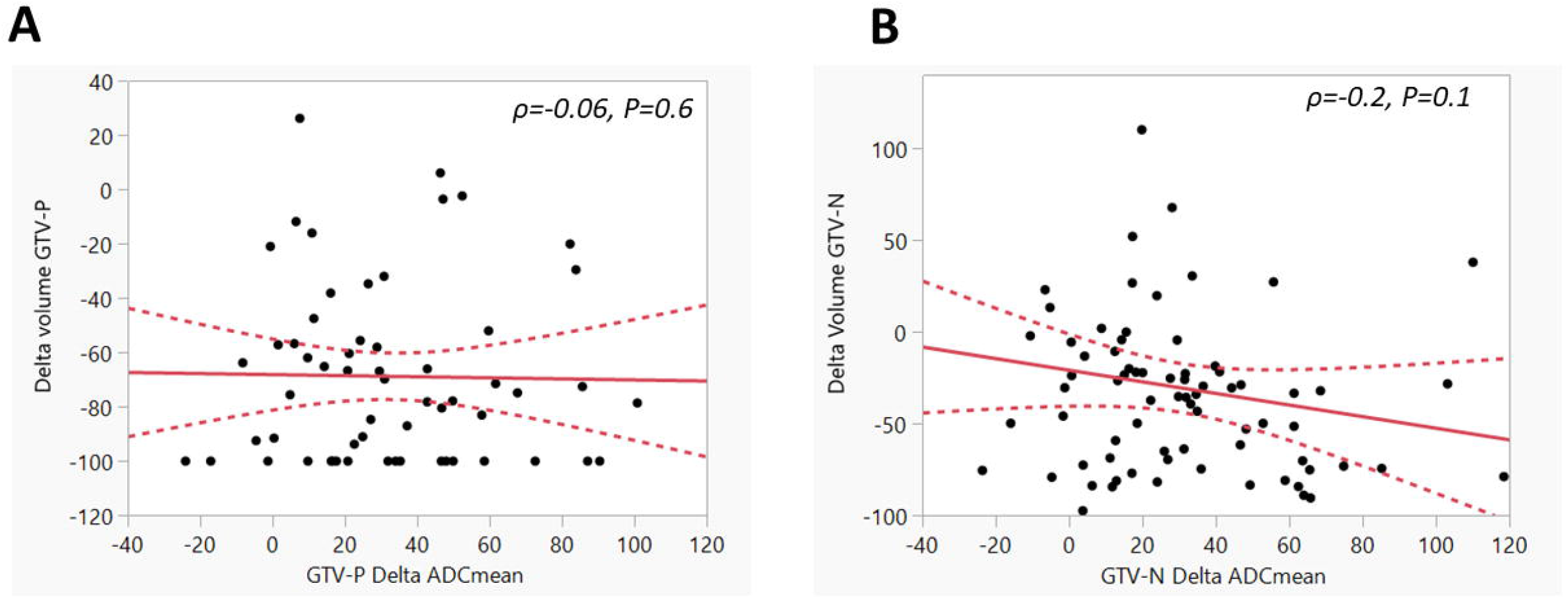
Relationship between Δ volume and ΔADC_mean_ for both GTV-P (A) and GTV-N (B) at mid-RT. Solid lines represent the linear fit and dotted lines represent the 95% confidence intervals.

#### ROI subvolume analysis

For patients with mid-RT non-CR at the primary site, there was no statistically significant difference in all ADC parameters between GTV-P-RS and GTV-P-RD (*p* >0.05 for all). RPA identified ΔADC_mean_ <5% and <10% as the strongest predictors of local recurrence for GTV-P-RD and GTV-P-RS, respectively (*p* =0.02 for both). However, for RFS only ΔADC_mean_ <5% for GTV-P-RD was significantly associated with worse RFS (*p* =0.01).

## Discussion

Our results show that DWI changes during RT are a significant predictor of oncologic outcomes. The significant increase in mid-RT ADC parameters for both tumor and nodal ROIs reflects a decrease in cellular density in tumor tissue caused by the radiation effect that induces breakdown of cellular membranes which ultimately decrease the restriction of diffusion shown in baseline tumor tissue.(33-35) The increased diffusion in mid-RT images was successfully measured by the studied ADC parameters that showed a higher increase in patients who ultimately developed CR at the end of treatment compared to patients with residual disease.

Our study also identified an ADC biomarker of local control and recurrence-free survival using a Δ GTV-P ADC_mean_ threshold of 7% increase relative to baseline ADC_mean_. These delta ADC changes were volume independent as our analysis methodology, illustrated in Figure 1, ensured that we use the same 3-D shape and volume of GTV-P propagated from baseline DWIs after image co-registration. We also assessed the effect of subvolume analysis within the subset of patients with non-CR at mid-RT images. In that subset, both ΔADC_mean_ changes in the residual and responding subvolumes were significantly associated with local control with a 5% and 10% threshold of ADC_mean_ increase. The threshold is lower in residual volumes as expected because of the higher relative tissue density in these subvolumes. This also indicates that quantitative DWI parameter maps can detect the mesoscale cellular changes that could not be otherwise detected using gross visual assessment. Furthermore, this also shows that even within the apparent residual tumors on anatomic imaging at mid-RT, there is a subset that expresses higher ADC changes and those tend to have better LC and RFS. These changes during treatment can serve as a biomarker to predict outcomes and can also be used as a biological tool to adapt therapy dose according to the predicted response during therapy.

An additional significant finding in our study is that pretreatment DWI parameters had no significant association with outcomes, indicating that dynamic information obtained from RT-induced imaging changes during treatment is likely more informative compared to baseline status. Several previous studies matched our findings of no association between pretreatment ADC and outcomes (20, 36, 37) while a prior pilot set from our group as well as other studies showed a significant correlation.(14, 38-40) We believe that pretreatment ADC parameters of a relatively homogenous cohort with a majority of HPV positive oropharyngeal cancer would be less predictive of outcomes when compared to a more heterogenous group of HNC subsites and/or tumor types. A heterogenous group of tumors will likely have a mixture of well and poorly differentiated tumors with different level of cellularity and stromal contents which thereby lead to more contrast in the degree of diffusion between different tumor types.(35) Therefore, we think that the pretreatment DWI parameters may be a more prognostic than predictive biomarker as it reflects the nature of the baseline tumor rather than predicting its response to therapy.(41)

In-treatment ΔADC were investigated in prior studies with relatively small sample sizes consisting of mixtures of HNC subsites, and in concordance with our results, these studies showed that ΔADC during RT was a significant correlate of oncologic outcomes.(11, 18, 36) To our knowledge, we present the largest prospective imaging study to date supporting that Δ primary tumor ADC changes during treatment are a strong biomarker of important oncologic outcomes, particularly for local control and recurrence-free survival. The threshold of ΔADC used should be carefully interpreted according to the nature of the primary tumor subsite, technique of segmentation/image registration, and DWI acquisition parameters (i.e., b values). Notably, ΔADC is a relative rather than an absolute value which could represent a more robust biomarker that is less susceptible to inter-patient and inter-scanner variability and thereby more clinically generalizable. In patients with mainly HPV positive oropharyngeal primary site using 3-D volumetric analysis of GTV-P at mid-RT relative to baseline, ΔADC_mean_ <7% was shown to be a strong correlate of local failure.

According to the criteria set by the Biomarker Qualification Program (BQP) that was developed by the Center for Drug Evaluation and Research (CDER, US Food and Drug Administration), a qualified biomarker must be within a specified context of use (COU) that defines the BEST biomarker category and its intended use.(41-43) Our findings from this prospective observational imaging study suggest that the COU for ΔADC_mean_ is as a response biomarker for defining HNC patients with high-risk of local failure according to their response to RT at an actionable mid-therapy time point. These results encourage us to apply for further full qualification package from the BQP with a properly defined COU. This will also allow a promising imaging biomarker like DWI to cross the second translational gap and eventually become a clinical decision-making tool according to the framework recommended by the imaging biomarker roadmap for cancer studies.(44)

However, our study is not without limitations. Importantly, our study utilized a single-institution cohort without external validation of our findings with multi-institutional data. Another limitation was the use of two DWI sequences during the study (i.e., BLADE and RESOLVE); however, after analyzing the ADC values extracted from both DWI sequences using the Kolmogorov–Smirnov test, no significant differences were found between the two sequences. Lastly, we failed to show any significant correlation between Δ nodal DWI changes and regional control, which could potentially be attributed to the cystic nature of the studied GTV-Ns in our sample. As a future step, we plan to analyze these LNs using a morphologic distinction between solid and cystic component in each node rather than the standard segmentation approach.

In conclusion, our prospective imaging study of HNC patients demonstrated that ΔADC parameters at mid-RT represent a strong predictor of local recurrence and recurrence-free survival. Patients with no significant increase of mid-RT ADC at the primary tumor site relative to baseline values are at high-risk of disease relapse. Multi-institutional data are needed for validation of our results.

## Data Availability

All data produced in the present study are available upon reasonable request to the authors.

